# Unrecognised Burden: High Prevalence and Clinical Impact of Fibromyalgia in Functional Motor Disorder

**DOI:** 10.1101/2025.07.02.25330711

**Authors:** Tereza Serranová, Lucia Nováková, Martin Jirásek, Barbora Křupková, Evžen Růžička, Michele Tinazzi, Tomáš Sieger

## Abstract

**Background:** Fibromyalgia is a chronic pain disorder, affecting 2-3% of the population, characterised by widespread pain, fatigue, sleep and cognitive symptoms. Despite symptom overlap between functional motor disorder (FMD) and fibromyalgia, the prevalence of fibromyalgia in FMD and its impact on health-related quality of life (HRQoL) remain unclear.

**Objectives:** To assess the prevalence of fibromyalgia in FMD using the 2016 American College of Rheumatology diagnostic criteria and to evaluate its impact on HRQoL.

**Methods:** A total of 139 consecutive patients with clinically established FMD (115 females, mean age 44.6 (SD 11.3) years) completed Fibromyalgia Survey Questionnaire, subjective motor symptoms and HRQoL assessments. Motor symptoms were objectively rated using the Simplified FMD Rating Scale (S-FMDRS). Major physical illnesses, neurological, and psychiatric comorbidities, and the use of centrally acting and nociceptive pain medication was recorded.

**Results:** Fibromyalgia was present in 44.6% of FMD patients (95%CI: 36.2–53.3%). Those with fibromyalgia had higher S-FMDRS scores (*p* < 0.01), lower HRQoL (*p* < 0.001), more frequent use of centrally acting and nociceptive pain medication (*p* < 0.01). Fibromyalgia severity was positively correlated with both subjective motor symptom severity (*p* < 0.01) and S-FMDRS (*p <* 0.05). Higher fibromyalgia severity (*p* < 0.001), S-FMDRS (*p* < 0.001), and psychiatric comorbidity (*p* < 0.001) were independent predictors of lower HRQoL.

**Conclusions:** In this study, fibromyalgia was common in FMD and associated with more severe motor symptoms, greater medication use and reduced quality of life, highlighting the importance of recognising and managing fibromyalgia in FMD.

---

Functional motor disorder (FMD) is a common source of neurological disability [1]. Its monetary impact is among the top healthcare-related expenditures for chronic illnesses [2]. People with FMD almost invariably present with numerous non-motor symptoms, such as pain, fatigue, and cognitive and psychological symptoms, which have a significant impact on health-related quality of life (HRQoL) [3]. Pain is one of the key symptoms of FMD [4,5]. Pain has been linked to greater FMD severity and reduced quality of life [6,7]. For instance, the presence of significant pain can prevent people with FMD from involvement in physical therapy. Assessing and treating pain is increasingly recognised as an integral part of managing FMD [8,9].

Fibromyalgia is a common chronic pain condition characterised by widespread pain that affects between 2-3% of the general population and has a substantial impact on people’s lives [10,11]. Fibromyalgia and FMD share common non-motor symptoms, including fatigue, cognitive issues, sleep disturbances, and headaches [7,12–14]. The importance of these non-motor symptoms has been recognised by the current diagnostic criteria, which abandoned the objective assessment of tender points, and the diagnosis is based on self-reported widespread pain and other symptoms [10,15].

According to a recent systematic review on pain in functional neurological disorder, the prevalence of fibromyalgia in different cohorts of people with functional neurological disorder has been reported to range from 4% to 18% across multiple studies, with a pooled prevalence of approximately 10% [5]. This rate is thus higher than estimates for the general population. However, it is likely still underestimated as the reported cases were not based on clinical assessment using the current criteria for fibromyalgia and establishing fibromyalgia was not the focus of those studies [16–21]. Given that fibromyalgia is frequently unrecognised and underdiagnosed, we hypothesised its true prevalence in FMD may be even higher and associated with a poorer HRQoL, more severe motor symptoms, higher multimorbidity burden, and increased medication use.

To address this, we systematically assessed the prevalence of fibromyalgia in a consecutive cohort of patients with FMD using the current diagnostic criteria. In addition, we collected detailed data on motor symptoms severity, neurological and systemic comorbidities, medication use, HRQoL. We then compared patients with and without fibromyalgia to explore the clinical correlates of comorbid fibromyalgia in FMD. Finally, we examined the association between the presence and the severity of fibromyalgia symptoms and HRQoL, to determine whether fibromyalgia independently predicts poorer quality of life in this population.

## Materials and methods

We recruited 163 consecutive patients (133 females, mean age 44.3 [standard deviation, SD=11.3, range 15 to 72] years; mean disease duration 6.9 [SD 7.3, range 0 to 40]) years with clinically definite FMD according to Gupta and Lang criteria between June 27, 2022, and February 23, 2024, from the specialized outpatient service for FMD at the Neurology Department of the First Faculty of Medicine and General University Hospital [22, 23]. The diagnosis of FMD was based on detailed clinical interviews and examination by a neurologist with experience with FMD (TS, LN) based on positive signs of internal inconsistency of the movement within one task or in between tasks [24].

Exclusion criteria were defined as: age <18 years, inability to complete questionnaires because of language difficulties, severe learning disabilities or cognitive impairment, major psychiatric comorbidity (e.g., psychosis, substance abuse). Comorbid neurological or rheumatological conditions were not exclusion criteria for the study participation.

### Objective assessment of motor symptoms

The motor symptoms were classified as functional weakness, tremor, dystonia, myoclonus, gait disorder, or speech disorder. Dominant (most severe and/or most frequent motor symptom) and additional motor symptom types (i.e. tremor, dystonia, gait disorder, myoclonus, and weakness) were identified. Each patient’s number of different motor symptoms was used as a proxy measure for motor disorder complexity.

The severity of the motor disorder was assessed using The Simplified Functional Movement Disorder Rating Scale (S-FMDRS) [25]. The presence or absence of abnormal movement at each of the seven body regions was recorded and rated according to symptom severity and duration (maximum score: 54). Gait aid score (10m minimal distance) was evaluated as normal gait = 0, abnormal gait no need for assistance or walking aids = 1, assistance or walker or crutches needed = 2, wheelchair dependent = 3) [7].

### Fibromyalgia diagnosis

The 2016 Fibromyalgia Survey Questionnaire was used to assess fibromyalgia based on the 2016 revisions to the 2010/2011 American College of Rheumatology (ACR) diagnostic criteria [15,26]. Fibromyalgia Survey Questionnaire evaluates the Widespread Pain Index (WPI) and Symptom Severity Scale (SSS), yielding a Fibromyalgia Severity Score (FSS). The WPI ranges from 0 to 19. The SSS score evaluates the severity of fatigue, unrefreshing sleep, cognitive symptoms, headaches, abdominal pain or cramps, and depression and ranges from 0 to 12. To meet the criteria for fibromyalgia (FM) diagnosis, an individual must have a WPI of 7 or higher and a SSS score of 5 or higher, or if the WPI is between 4 and 6, the SSS score must be 9 or higher [15]. In addition, the individual must experience generalised pain, defined as pain in at least four of five regions, with symptoms persisting for at least three months. To ensure accuracy, a back-translated version of the questionnaire was developed with input from bilingual experts and subject-matter specialists.

### Subjective assessment of motor symptoms severity and HRQoL

All patients self-evaluated their motor symptom severity on a 3-point Likert scale (not bothered at all = 0, bothered a little = 1, bothered a lot = 2) according to the adjusted Patient-Health Questionnaire (PHQ-15) [27]. In addition to PHQ-15 items assessing weakness, motor coordination impairment and gait disorder, we added one item assessing tremor and jerks and one item assessing abnormal postures or spasms. The total score (subjective motor symptoms severity, SMSS, range 0-10) was calculated [7].

HRQoL was assessed using the 12-Item v2 Short Form Health Survey (SF-12) [28]. Physical Functioning, Role Limitations (both Physical and Emotional), Social Functioning, Pain, Mental Health, Vitality and General Health are HRQoL domains reflected in SF-12 (total score 12-44).

### Clinical assessment of comorbidities and medication

All patients were interviewed for the presence of typical symptoms of migraine and restless legs syndrome, and the current diagnostic criteria were applied to establish the diagnoses. The presence of comorbidities including: neurological comorbidities (radicular pain, polyneuropathy, infectious disease, essential tremor, Parkinson’s disease, dystonia, stroke, demyelinating disease, neuromuscular disorder, epilepsy or other), physical health comorbidities (arterial hypertension, diabetes mellitus, cardiovascular disease, dyslipidaemia, thyroid disorders, asthma, gastrointestinal disorder, rheumatic disease, autoimmune disease, gynaecological and reproductive disorders, history of malignancy, obesity) was recorded based on reliable medical report review. Given the absence of systematic psychiatric assessments in all patients, the presence of psychiatric comorbidity was recorded dichotomously (present/absent), based on medical documentation, clinical history, screening during neurological examination, and responses to self-report questionnaires assessing depression and anxiety.

The use of centrally acting and nociceptive medications including gabapentin, pregabalin, selective serotonin reuptake inhibitors (SSRIs), serotonin-norepinephrine reuptake inhibitors (SNRI) e.g., venlafaxine, serotonin antagonist and reuptake inhibitors (SARI), noradrenergic and specific serotonergic antidepressants (NaSAs), tricyclic antidepressants (TCAs), bupropion, dopamine agonists, antihistamines, dopamine antagonists, benzodiazepines, opioids, anticonvulsants, cyclobenzaprine, nonsteroidal anti-inflammatory drugs (NSAIDs), paracetamol, triptans and medicinal cannabis, was recorded for all participants.

Statistics: The Fisher test was used to test independence in contingency tables. The Wilcoxon exact rank test was used to test group differences in continuous and discrete measures [29]. Pearson correlation was used to study the relationship between WPI and motor symptom severity measures. ANOVA was used to test how FSS differed between patients who used different gait aids. Poisson regression was used to study how the number of motor phenotypes was related to the FSS. A linear model was used to find out whether SF-12 could be explained using the combination of FSS, S-FMDRS, lifelong psychiatric comorbidity, multimorbidity, overall medication use, age, and sex. The Holm method was used to correct for the number of tests performed within categories (motor phenotypes, motor symptoms measures, medication, and comorbidities) [30].

Ethical consideration: The study complies with the Declaration of Helsinki’s ethical standards. The study was approved by the local ethics committee, and all participants gave their written consent to take part in the study 53/23 Grant AZV VES 2024.

## Results

In 24 out the 163 patients recruited, key data were missing (SF-12 in 12 of them, QPC in 15, SMSS, FPH1 and STAI.X2 in 10, etc.), so we restricted our sample to 139 patients with FMD (115 females, mean age 44.6 [standard deviation, SD=11.3, range 15 to 72] years; mean disease duration: 6.9 [SD 7.3, range 0 to 40] years).

### Prevalence of fibromyalgia in FMD

In the analysed sample of 139 FMD patients, 62 patients fulfilled the diagnostic criteria for fibromyalgia, i.e. the prevalence of fibromyalgia was 44.6% (95% CI (36.2, 53.3) %).

Patients with fibromyalgia (FM+) and patients without fibromyalgia (FM-) did not differ significantly in age, sex, age at disease onset, or disease duration (see Table 1 for details).

**Table 1.**
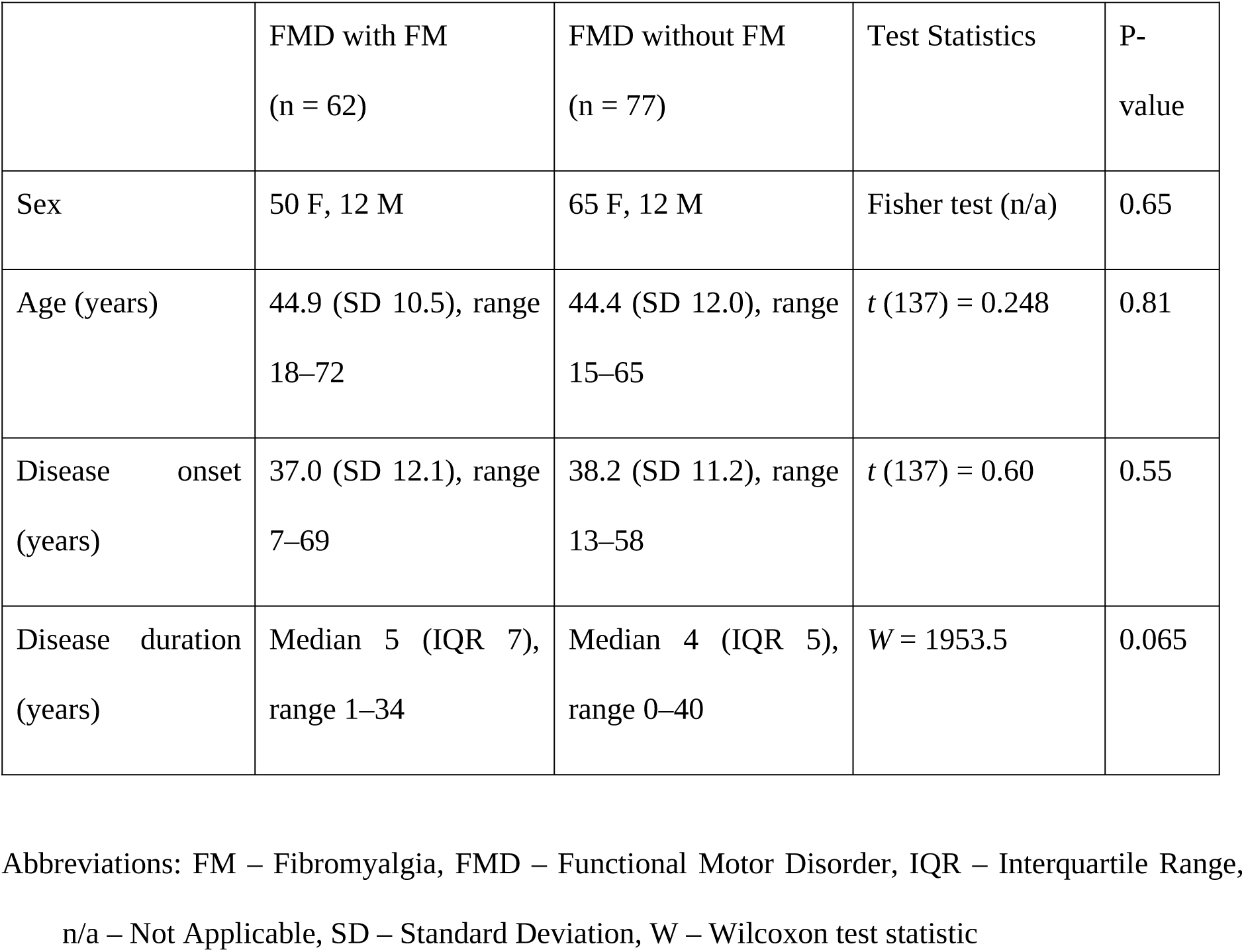
Demographic and clinical characteristics in FMD patients with and without fibromyalgia.

### Between group comparison of FM+ versus FM-patients

FM+ patients had decreased HRQoL compared to those without FM (median values 20 vs. 24, range 12 to 32 vs. 14 to 42, W=3480, *p* < 0.001) *(also in Table 2)*.

**Table 2.** Health-related quality of live and motor symptom measures in FMD patients with and without fibromyalgia.

|  | FMD with FM<br>(n = 62) | FMD without<br>FM (n = 77) | Test Statistics | P-value<br>(corrected) |
| --- | --- | --- | --- | --- |
| **SF-12 (median [IQR], range)** | 20 [6], 12–32 | 24 [6], 14–42 | W = 3480 | **< 0.001** |
| **Main motor phenotype (n)** |  |  |  |  |
| Tremor | 17 | 22 | Fisher test (n/a) | 1 |
| Gait disorder and instability | 23 | 13 | Fisher test (n/a) | 0.065 |
| Dystonia | 7 | 8 | Fisher test (n/a) | 1 |
| Weakness | 13 | 32 | Fisher test (n/a) | 0.065 |
| Myoclonus | 2 | 1 | Fisher test (n/a) | 1 |
| Speech and swallowing<br>disorder | 0 | 1 | Fisher test (n/a) | 1 |
| Parkinsonism | 0 | 0 | n/a | n/a |
| **Main or additional motor phenotype (n)** |  |  |  |  |
| Tremor | 38 | 42 | Fisher test (n/a) | 1 |
| Gait disorder and instability | 49 | 47 | Fisher test (n/a) | 0.19 |
| Dystonia | 23 | 20 | Fisher test (n/a) | 1 |
| Weakness | 47 | 52 | Fisher test (n/a) | 1 |
| Myoclonus | 7 | 4 | Fisher test (n/a) | 1 |
| Speech and swallowing | 7 | 9 | Fisher test (n/a) | 1 |
| disorder |  |  |  |  |
| Parkinsonism | 1 | 0 | Fisher test (n/a) | 1 |
| **S-FMDRS score (median, range)** | 14.5 (0–34) | 10 (0–31) | W = 1707 | **0.016** |
| **Gait disturbances (%)** | 77% | 56% | Fisher test (n/a) | **0.026** |
| **Use of gait aids (%)** | Wheelchair:<br>8%<br>Crutch: 26% | Wheelchair:<br>3%, Crutch:<br>13% | W = 2832.5 | **0.026** |
| **No. of motor phenotypes (median [IQR], range)** | 3 [1], (1–6) | 2 [1], (1–6) | W = 1796.5 | **0.026** |
| **SMSS (median [IQR], range)** | 7.5 [3], 1–10 | 5 [4], 0–10 | W = 1230.5 | **< 0.001** |

Abbreviations: FM – Fibromyalgia, FMD – Functional Motor Disorder, HRQoL – Health-Related
Quality of Life, IQR – Interquartile Range, n/a – Not Applicable, S-FMDRS –The Simplified
Functional Movement Disorder Rating Scale, SF-12 – 12-Item v2 Short Form Health Survey SMSS –
Subjective Motor Symptom Severity, W – Wilcoxon test statistic
Significant results are presented in bold.

While there was a significant association between the main motor phenotype and the presence of fibromyalgia (Fisher test *p* = 0.026), evaluation of individual phenotypes present either as main or additional did not reveal any significant associations with fibromyalgia after correction for multiple comparisons across the six phenotypes. FM+ patients showed higher SFMDRS scores ( *p* = 0.016), more frequent gait disturbances (*p* = 0.026), greater reliance on gait aids (*p* = 0.026), and a higher number of motor phenotypes (*p* = 0.026). Subjective motor symptom severity was also greater in FM+ (*p* < 0.001). Gait disorder and instability were relatively more common, and weakness less common, in FM+ patients (both *p* = 0.065), though overall phenotype frequencies were similar across groups (see Table 2 for details).

The total number of comorbid physical illnesses and neurological comorbidities, migraine, and RLS frequencies did not differ significantly between FM+ and FM-groups (all *p* > 0.05; see Table 3 for details and Supplementary Table 1 for Neurological comorbidities and Supplementary Table 2 for comorbid major physical illnesses).

**Table 3.** Medication and comorbidities in FMD patients with and without fibromyalgia.

|  | FMD with FM<br>(n = 62) | FMD without<br>FM<br>(n = 77) | Test Statistics | P-value<br>(corrected) |
| --- | --- | --- | --- | --- |
| Medication |  |  |  |  |
| Number of medications<br>(median [IQR], range) | 2 [2], 0-7 (most<br>frequent: 2<br>meds) | 1 [3], 0-8 (most<br>frequent: 0<br>meds) | W = 1712 | **0.012** |
| Use of FM-specific agents<br>(amitriptyline, cannabinoids,<br>tramadol) (%) | 58% | 32% | W = 1771 | **0.012** |
| Use of nociceptive painkillers<br>(NSAIDs, paracetamol) (%) | 66% | 38% | W = 1784.5 | **0.012** |
| Other medication<br>(antidepressants, opioids) | 47% | 53% | W = 2182.5 | 0.36 |
| Comorbidities |  |  |  |  |
| Total number of comorbidities<br>(median [IQR], range) | 3 [2], 0-9 | 3 [2], 0-7 | W = 1926 | 0.24 |
| Number of internal<br>comorbidities (median [IQR],<br>range) | 2 [2], 0-7 | 1 [1], 0-5 | W = 2039.5 | 0.38 |
| Number of neurological | 0 [1], 0-2 | 0 [0], 0-2 | W = 2243 | 0.85 |
| comorbidities (median [IQR], range) |  |  |  |  |
| Migraine prevalence (%) | 55% | 39% | Fisher test (n/a) | 0.35 |
| RLS prevalence (%) | 37% | 38% | Fisher test (n/a) | 1 |

Abbreviations: FM – Fibromyalgia, FMD – Functional Motor Disorder, IQR – Interquartile Range,
n/a – Not Applicable, NSAIDs – Non-Steroidal Anti-Inflammatory Drugs, RLS – Restless Legs
Syndrome, W – Wilcoxon test statistic
Significant results are presented in bold.

FM+ patients used significantly more medications than FM– patients (*p* = 0.012). They more frequently received drugs with some efficacy in fibromyalgia (amitriptyline, cannabinoids, tramadol; *p* = 0.012) and nociceptive painkillers (*p* = 0.012), while no group differences were observed for other drug classes, including antidepressants, benzodiazepines, opioids (see Table 3 for details).

### Correlation analysis of FSS with objective and subjective motor symptom severity measures

Objective measures: S-FMDRS correlated with FSS (the sum of WPI+SS score) *(r* = 0.20, *t* (137) = 2.38, *p* = 0.019). On average, the number of motor phenotypes was greater for higher FSS (Poisson regression *χ^2^*(1) = 4.12, *p* = 0.04). FSS differed between patients using different gait aids (those using no gait aids, the mean FSS was 14.9 (range 1 to 30), in those using crutch 17.9 (range 5 to 31), and in those using a wheelchair 18.3 (range (16 to 22) (ANOVA *F* (1,137) = 5.0, *p* = 0.027). There was a strong relation between subjective assessment of motor symptom severity, i.e. SMSS and FSS both in FM-patients (*r* = 0.71, *t* (75) = 8.58, *p* <0.001) and in FM+ patients (*r* = 0.34, *t* (60) = 2.85, *p* = 0.006).

### Predictors of the HRQoL

SF-12 could be partially explained by the combined effect of FSS, S-FMDRS, and lifelong psychiatric comorbidity (an increase of each of them was, on average, associated with worsening of HRQoL, FSS: an increase of 10 points was associated with SF-12 worsening by 4.5 points, *t* (135) = −8.03, *p* < 0.001; S-FMDRS: an increase of 10 points was associated with SF-12 worsening by 1.6 points, *t* (135) = −3.62, *p* < 0.001; psychiatric comorbidity: their presence resulted in decrease of SF-12 by 3.2, *t* (135) = −4.40, *p* < 0.001; adjusted *R^2^* = 0.47).

## Discussion

This is the first study to apply the 2016 ACR criteria in a well-characterised consecutive sample of patients with FMD, revealing a very high prevalence of comorbid fibromyalgia. The presence of fibromyalgia was associated with lower HRQoL and worse subjectively and objectively rated motor symptom severity and increased use of medication with central effect and medication used for nociceptive pain.

With 44.6% of FMD patients meeting the criteria for fibromyalgia, we found a prevalence at least ten times higher than that reported in the general population [10]. The prevalence of fibromyalgia diagnosed using the current diagnostic criteria in this study was also much higher than in the studies that reported fibromyalgia cases in various cohorts of patients with functional neurological disorders [16–20]. The frequency of previously reported fibromyalgia diagnoses varied from 7/176 (4%) patients in FMD cohorts to 6/56 (16%) [16–20]. These studies, however, did not systematically assess fibromyalgia using the current diagnostic criteria, relying instead on retrospective chart reviews. This likely contributed to underrecognition of the condition, highlighting that unless standardised criteria are actively applied, fibromyalgia may remain frequently overlooked [31].

High frequency of fibromyalgia using the current criteria has been reported in numerous conditions, including other somatic symptom disorders such as chronic fatigue syndrome [32]; other pain conditions, including migraine [33–35] or disorders associated with nociceptive pain, such as rheumatoid arthritis [33]. In our cohort, however, the multimorbidity, including neurological conditions (including migraine and RLS), did not differ between patients with and without fibromyalgia, and we did not find an association between multimorbidity and fibromyalgia severity. This finding suggests that fibromyalgia is not primarily associated or driven by comorbidities.

Available epidemiological data suggest that FM (2-8% in the general population) is more prevalent than functional neurological disorder (80-140 per 100,000) [36]. Although many people seen in chronic pain services have a history of functional neurological disorder (17%) [37], there is a lack of knowledge of the prevalence of FMD in fibromyalgia. While people with fibromyalgia present with increased frequency of neurologic signs and symptoms, including motor abnormalities, specific tests for establishing FMD (i.e. tests demonstrating inconsistencies such as Hoover’s sign or distractibility) were not performed [38].

FMD patients with fibromyalgia presented with more severe motor symptoms measured objectively using the S-FMDRS and a higher number of motor phenotypes. Patients with fibromyalgia had a greater frequency of gait disturbances and increased reliance on gait aids such as wheelchairs and crutches. Significant difference was also found in subjectively reported motor symptom severity, which was higher in the fibromyalgia group. These findings align with previous observations from other studies, which found that severity in motor and non-motor symptoms is highly cross-correlated in FMD patients and further support a hypothesised shared mechanism of the two conditions [7,39]. Several studies reported a lack of association of a particular phenotype and non-motor symptoms profile [7,40]. Similarly, there was no significant association of fibromyalgia with a specific motor phenotype in this study. The findings from this study suggest that the presence of fibromyalgia should be systematically assessed in all FMD patients, particularly those with more severe motor involvement.

We found a significant impact of fibromyalgia on HRQoL. FMD patients with fibromyalgia reported lower HRQoL, even after correction for depression/anxiety. This finding is in line with previous reports on the impact of pain and other non-motor symptoms on HRQoL and it highlights the importance of addressing them in the management [3,7] As the presence of self-reported non-motor symptoms is part of the current diagnostic criteria for fibromyalgia, we did not investigate symptoms such as fatigue, depression, anxiety or sleep separately. There was a positive association between fibromyalgia severity and subjectively and objectively assessed motor symptoms. However, we found no significant difference in multimorbidity between FMD patients with and without fibromyalgia, suggesting that the presence of fibromyalgia is not primarily driven by other common comorbid conditions.

Patients with fibromyalgia used a greater number of centrally acting and pain medications compared to patients without fibromyalgia. While a significant proportion of patients with fibromyalgia (58 %) was on medication with some evidence of efficacy in fibromyalgia, such as amitriptyline, cannabinoids, and tramadol, even a larger proportion of patients with fibromyalgia (66%) reported using nociceptive painkillers (non-steroidal analgesics and paracetamol) with limited/no evidence of efficacy. Our findings are in line with previously published reports on pain medication in motor and seizure subtypes of FND, which reported up to 50% of individuals on pain medication [17,41,42]. The increased use of nociceptive pain medications may reflect frequent misdiagnosis and focus on local pain problems and limited recognition of the nociplastic nature of chronic pain conditions, as well as insufficient patient education regarding chronic nociplastic pain and non-pharmacological treatment options. Additional contributing factors may include cultural beliefs, lack of education of health care professionals, the absence of national treatment guidelines for fibromyalgia, and the off-label status of effective medications.

Our study highlights the importance of widespread pain assessment and its adequate management in the FMD population. Pain is associated with greater FND severity, reduced quality of life, and worse prognosis [5,7,20,43]. Diagnosis of fibromyalgia in FMD has numerous implications for the management, including provision of pain education and adjustment of self-management plan, which should focus on maintaining activity, not symptom relief, lower-grade exercise, and triage for specialised pain care [10]. FSS is a reliable negative prognostic factor for outcomes for several types of interventions, including hip surgery or persistent pelvic pain following hysterectomy [44,45]. These findings can also apply to people with FMD and guide clinical decision making and planning, such as avoiding overmedication, consulting multiple specialists and unnecessary surgeries for local pain problems in people with fibromyalgia through multidisciplinary care. There are international guidelines and other evidence-based recommendations for pharmacological and non-pharmacological management of fibromyalgia, including education, physical activity/rehabilitation techniques and other non-invasive treatments interventions based on comorbidities such as mood and sleep disorders, which can also be used for patients with FMD in the absence of specific recommendations [46–48]. Treatments potentially effective for pain, such as physical therapy and behavioural interventions, mostly failed to improve pain in trials with people with FMD [5,49–51]. One recent study combining physiotherapy and cognitive behavioural therapy reported improvement in pain [52]. Identifying FMD patients with comorbid symptoms such as fibromyalgia could help guide clinicians toward recommending brain–mind–body approaches, which may be more appropriate than standard motor rehabilitation alone.

Functional neurological disorder and other somatic syndrome disorders including fibromyalgia are associated with alterations in the functioning of brain networks and are now believed to share underlying pathophysiological mechanisms [39,53]. Recent theoretical models based on predictive coding accounts of brain function suggest functional symptoms arise from the development of abnormal predictions about motor and sensory states, including pain, driven by an abnormal allocation of attention [39,53]. These models are symptom type agnostic and suggest that a common underlying mechanism can explain motor, sensory, cognitive, and interoceptive manifestations. Despite growing clinical and scientific evidence for a unified mechanism, pain, fatigue, and other non-motor symptoms in FND remain separately classified in DSM-5 as somatic symptom disorder and chronic pain syndrome [23]. A diagnostic split also exists in the 11th revision of the International Statistical Classification of Diseases and Related Health Problems (ICD-11), where FMD appears in both the Neurology (08) and Psychiatry chapters (06), while chronic widespread pain is coded separately under Symptoms, signs or clinical findings, not elsewhere classified (21) [54]. The high prevalence of fibromyalgia in FMD further challenges this classification divide and underscores the need for more integrated diagnostic frameworks.

There are limitations to this study, including bias inherent to a single-centre epidemiological study. People with more severe FMD are likely to be followed in a tertiary centre with specialised FMD service, which results in a potential overestimation of fibromyalgia frequency in the FMD cohort derived from such a centre. We did not assess whether fibromyalgia developed before or after the onset of motor symptoms, nor the time interval between the two. Additionally, we did not systematically ask patients whether they had undergone a rheumatological assessment. These limitations should be addressed in future studies. A multicenter study from different countries would increase sample size and diversity, reduce selection bias, provide a more representative sample of the FMD population, and allow for examining cross-cultural differences. A recent review on pain in FND suggested that pain is more common in FMD than in functional seizures [5]. Future studies should assess the prevalence of fibromyalgia in functional seizures.

## Conclusions

Fibromyalgia was highly prevalent in FMD and was associated with more severe motor symptoms, higher medication use, and lower quality of life. This finding may have implications for the assessment and management of FMD patients with comorbid pain. Systematic screening for fibromyalgia may be warranted in FMD clinics, particularly in specialized centres, to ensure appropriate multidisciplinary care and tailored treatment strategies.

## Data Availability

The data analysed in the present study are available upon reasonable request.

## Acknowledgements

Funding: This study was supported by the Czech Ministry of Health (NW24-04-00456 and MH CZ– DRO-VFN00064165) and National Institute for Neurological Research (Programme EXCELES, ID Project No. LX22NPO5107), and the Czech Ministry of Education, Youth and Sports /EU ERDF-Project Brain Dynamics, No. CZ.02.01.01/00/22_008/0004643.

## Author roles

1. Research project: A. Conception, B. Organization, C. Execution;
2. Statistical Analysis: A. Design, B. Execution, C. Review and Critique;
3. Manuscript: A. Writing of the first draft, B. Review and Critique;
4. Funding: A. Obtaining Funding.

**Tereza Serranová:** 1A, 1B, 1C, 2A, 2C, 3A, 3B, 4A

**Lucia Nováková:** 1A, 1B, 1C

**Martin Jirásek:** 1C, 2B, 3A

**Barbora Křupková:** 1C, 2B, 3A

**Evžen Růžička:** 3A, 3B

**Michele Tinazzi:** 3A, 3B

**Tomáš Sieger:** 2A, 2B, 2C, 3A

## Disclosures

Tereza Serranová

**Stock Ownership in medically-related fields:** None

**Intellectual Property Rights:** None

**Consultancies:** None Expert Testimony: None

**Advisory Boards:** None

**Employment:** Department of Neurology and Centre of Clinical Neuroscience, Charles University in Prague, 1^st^ Faculty of Medicine and General University Hospital in Prague, Czech Republic

**Partnerships:** None

**Contracts:** None

**Honoraria:** None

**Royalties:** None

**Grants:** Ministry of Health of the Czech Republic

**Other:** None

**Lucia Nováková**

**Stock Ownership in medically-related fields:** None

**Intellectual Property Rights:** None

**Consultancies:** None Expert Testimony: None

**Advisory Boards:** None

**Employment:** Department of Neurology and Centre of Clinical Neuroscience, Charles University in Prague, 1^st^ Faculty of Medicine and General University Hospital in Prague, Czech Republic

**Partnerships:** None

**Contracts:** None

**Honoraria:** None

**Royalties:** None

**Grants:** Ministry of Health of the Czech Republic

**Other:** None

**Martin Jirásek**

**Stock Ownership in medically-related fields:** None

**Intellectual Property Rights:** None

**Consultancies:** None Expert Testimony: None

**Advisory Boards:** None

**Employment:** Department of Department of Rehabilitation and Sports Medicine, Charles University in Prague, 2^nd^ Faculty of Medicine and University Hospital Motol in Prague, Czech Republic

**Partnerships:** None

**Contracts:** None

**Honoraria:** None

**Royalties:** None

**Grants:** Ministry of Health of the Czech Republic

**Other:** None

**Barbora Křupková**

**Stock Ownership in medically-related fields:** None

**Intellectual Property Rights:** None

**Consultancies:** None Expert Testimony: None

**Advisory Boards:** None

**Employment:** Department of Neurology and Centre of Clinical Neuroscience, Charles University in Prague, 1^st^ Faculty of Medicine and General University Hospital in Prague, Czech Republic

**Partnerships:** None

**Contracts:** None

**Honoraria:** None

**Royalties:** None

**Grants:** Ministry of Health of the Czech Republic

**Other:** None

**Evžen Růžička**

**Stock Ownership in medically-related fields:** None

**Intellectual Property Rights:** None

**Consultancies:** None Expert Testimony: None

**Advisory Boards:** None

**Employment:** Department of Neurology and Centre of Clinical Neuroscience, Charles University in Prague, 1^st^ Faculty of Medicine and General University Hospital in Prague, Czech Republic

**Partnerships:** None

**Contracts:** None

**Honoraria:** None

**Royalties:** None

**Grants**: Ministry of Health of the Czech Republic

**Other:** None

**Michele Tinazzi**

**Stock Ownership in medically-related fields:** None

**Intellectual Property Rights:** None

**Consultancies:** None Expert Testimony: None

**Advisory Boards:** None

**Employment:** Department of Neurosciences, Biomedicine and Movement Sciences, University of Verona, Verona, Italy

**Partnerships:** None

**Contracts:** None

**Honoraria:** personal fees from Abbvie, Bial, Chiesi Farmaceutici, Zambon, and the Movement Disorders Society.

**Royalties:** None

**Grants**: Italian Ministry of Health, University of Verona, and the Verona Brain Research Foundation

**Other:** None

**Tomáš Sieger**

**Stock Ownership in medically-related fields:** None

**Intellectual Property Rights:** None

**Consultancies:** None Expert Testimony: None

**Advisory Boards:** None

**Employment:** Department of Cybernetics, Faculty of Electrical Engineering, Czech Technical University in Prague, Czech Republic and Department of Neurology and Centre of Clinical Neuroscience, Charles University in Prague, 1^st^ Faculty of Medicine and General University Hospital in Prague, Czech Republic

**Partnerships:** None

**Contracts:** None

**Honoraria:** None

**Royalties:** None

**Grants:** None

**Other:** None

## Supplementary Tables

**Table S1.** Frequencies of Neurological Comorbidities in FMD Patients (n = 139)

|  | n |
| --- | --- |
| Radicular pain | 10 |
| Stroke | 5 |
| Neuromuscular disorder | 5 |
| Traumatic brain injury | 5 |
| Polyneuropathy | 4 |
| Essential tremor | 3 |
| Cranial nerve disorder (i.e. facial nerve palsy) | 3 |
| Dystonia | 2 |
| Epilepsy | 2 |
| History of infectious disease of nervous system | 1 |
| Demyelinating disease | 1 |

**Table S2.** Frequencies of comorbid physical illnesses in FMD Patients (n = 139)

|  | n |
| --- | --- |
| Obesity | 44 |
| Arterial hypertension | 32 |
| Asthma or other respiratory disorder | 26 |
| Thyroid disorder | 25 |
| Rheumatological disorder | 21 |
| Dyslipidemia | 18 |
| Cardiovascular disease | 17 |
| Gastrointestinal disorder | 15 |
| Gynecological and reproductive disorders | 10 |
| Diabetes mellitus | 8 |
| History of malignancy | 5 |
| Autoimmune disorder | 4 |

## Notes

### Competing Interest Statement

The authors have declared no competing interest.

### Author Declarations

Ethics committee of the General University Hospital in Prague gave ethical approval for this work. The approval number is 53/23 Grant AZV VES 2024). All participants gave their written consent to take part in the study.

### Summary of Updates

This version of the manuscript has been revised to update the copyright options explicitly stated in the manuscript file to CC-BY.

